# Cost-Benefit Analysis of Far-UVC Lamps for Reducing Indoor Infection Transmission in Switzerland and Germany: Insights from the CERN Airborne Model for Indoor Risk Assessment (CAiMIRA)

**DOI:** 10.1101/2025.04.02.25325071

**Authors:** Sabine Matysik, Elina Christian, Bianca Bohmann, Laurent Bächler, Stefan Krüger, Marwan El Chamaa, Markus Baumeister, Sungmin Eu

**Author notes:** Corresponding author: Elina Christian < >. **Contact information** Matysik, Sabine < >, Laurent Bächler < >, “Bohmann, Bianca” < >, “El Chamaa, Marwan” < >, “Baumeister, Markus” < >, Daniel Staudenmann < >, Jasper Götting < >, “ ” < >, “Krueger, Stefan” < >, “Eu, Sungmin”. **JEL** I 18 (Government Policy • Regulation • Public Health), Q 53 (air pollution), Q 55 (technological innovation).

## Abstract

Far-UVC light (wavelengths 207-230 nm) can be used directly overhead, whilst having germicidal capabilities to improve indoor air quality. This study evaluates the cost-benefit of implementing far-UVC devices in various settings in Switzerland and Germany. We used the CERN Airborne Model for Indoor Risk Assessment (CAiMIRA) to model infection risk reduction in restaurants, offices, and waiting rooms, considering factors like room size, occupancy, and ventilation rates. Three scenarios were analysed: a normal winter (22 weeks), a COVID-19-like pandemic (4-week wave), and a severe pandemic (8-week wave). Avoided infections were translated into healthcare, economic and avoided quality-adjusted life years (QALY) metrics. Costs included purchasing, installing, maintaining, and operating UV-C lamps. In Switzerland, cost-benefit ratios ranged from one franc to: 30-290 CHF during a normal winter; 65-430 CHF during a COVID-like pandemic; and 2,300-20,500 CHF during a severe pandemic. In Germany, cost benefit ratios ranged from 1 euro to: 7-226 EUR during a normal winter; 118-449 EUR during a COVID-like pandemic; and 659-18,946 EUR during a severe pandemic. Far-UVC lamps are a highly cost-effective solution for societies during normal winter and pandemic scenarios. Implementation in the settings studied should be considered as a safe and effective measure for infectious disease control.

## Introduction

Ultraviolet light has been investigated as a disinfection technology for the past century, but use indoors has been limited to indirect, upper air disinfection due to excess UV radiation exposure concerns [1, 2, 3, 4]. Far-UVC light are wavelengths 207-230 nm that are of interest for the ability to be used directly overhead, whilst still having germicidal capabilities [5, 6]. Indoor environments are particularly conducive to the transmission of airborne pathogens, contributing to the propagation of infectious diseases [7]. For example, over 90% of SARS-CoV-2 infections occurred indoors [7]. Since most people spend the majority of their time indoors, technologies to improve indoor air quality are crucial to mitigate transmission of pathogens [8, 9]. Far-UVC light, which is wavelengths of light between 200 and 230 nm within the UV spectrum, is a promising approach for real-time infection mitigation [10]. Lights directly overhead busy indoor environments provide the opportunity to neutralise the threat of pathogens as they emerge from a contagious person. Duration of exposure to the UV lights needed to break down the pathogenic DNA or RNA depends on many factors.[1, 5, 11] This study seeks to address whether investments in mitigating indoor infections using far-UVC provide worthwhile returns in Switzerland. Previously, to our knowledge the cost-benefit ratio associated with the utilisation of far-UVC light had not been determined.

### Existing measures to enhance indoor air quality

Indoor air technologies provide effective mitigation of disease transmission, especially when used in conjunction with one another. The cornerstones of indoor air quality technology are:

⍰ Ventilation improves air quality by moving air, either naturally or mechanically. It is dependent on a reasonable frequency of air changes to reduce respiratory airborne particle accumulation.[12,13]
⍰ Filtration physically removes particulate matter and sometimes pathogens through different materials and processes.[14]
⍰ Upper air germicidal UV light is typically used indirectly in upper room levels or as part of an air conditioning system as the high wavelength is dangerous for human exposure.[3]

Far-UVC light adds to this repertoire as it can be installed overhead in occupied indoor spaces due its enhanced safety profile.

### Efficacy

Far-UVC light is effective at inactivating airborne viruses like influenza and SARS-CoV-2 by damaging their proteins and nucleic acids [11, 15, 16, 17]. Non-enveloped viruses, with robust capsid proteins, require a higher dose and longer exposure to achieve similar inactivation levels [18]. There is variation between types of pathogen and even strains, however consistent antimicrobial activity is demonstrated; it may require longer to reduce the pathogen load in the room [1, 10].

### Safety

The International Commission on Non-Ionizing Radiation Protection (ICNIRP) has recommended exposure limits for ultraviolet radiation that are designed to protect the skin and eyes. The ICNIRP recommends an exposure dosage limit for 222 nm far-UVC light of 240 J/m² (24 mJ/cm²), with an exposure time limit of 8 hours per day and 40 hours per week [4]. The ICNIRP exposure limits form the basis of the European standards [19]. The current European exposure limits are based on recommendations from the 1970s [20]. Based on safety studies from the last decade, the American Conference of Governmental Industrial Hygienists, in 2021, has markedly raised their recommendations [20, 21, 22].

There have been numerous studies evaluating the effects of far-UVC as a potential carcinogen, with results demonstrating absorption within the outermost layers of the skin and eyes, and no increased risk [1, 6, 23]. A key safety consideration for far-UVC is whether a filter to remove longer wavelengths of light is included [10]. There may be unknown longer-term effects of far-UVC exposure, for instance on the skin microbiome and immune system, which remain underexplored [24, 25].

Furthermore, far-UVC light can interact with atmospheric molecules like oxygen and volatile organic compounds, leading to the formation of ozone and harmful radicals indoors[26, 27, 28]. Directive 2008/50/EC mandates ozone concentration limits of less than 120 μg/m³ over an 8-hour average period [29]. Monitoring of ozone precursors like nitrogen oxides and volatile organic compounds is required to ensure air quality compliance [29]. Initial data shows that minimal ventilation is required to mitigate the effects of far-UVC on indoor particulate matter, although this is reliant upon a low external ozone concentration [30]. In brief, adherence to the ICNIRP limits for far-UVC usage in combination with ventilation appears reasonably practicable and safe based on current understanding.

### Biosecurity significance

The increasing risk from emerging infectious disease and recent experience during the COVID-19- pandemic underscore the importance of measures to reduce the risk of transmission in indoor settings [31]. Studies have demonstrated great variation in the efficacy of personal protective and environmental infection control measures [32, 35, 36]. Measures with lower level of personal restriction compared to effect on containment of the pandemic were associated with higher levels of public acceptance in Germany, suggesting that far-UVC could become a well-accepted pandemic response measure [33].

Slowing early transmission is a crucial first step in epidemic and pandemic response [34]. Yet, emerging or newly mutated pathogens often spread before relevant mitigation measures are implemented [34]. To give time for pathogen specific responses such as medication to be enacted, non-pharmaceutical interventions like restriction of movement, social distancing and mask wearing can be implemented, but are subject to individual compliance [34, 35]. Far-UVC may add to existing non-pharmaceutical intervention options by reducing pathogen load indoors in a manner that is not as disruptive as existing interventions. Furthermore, there can be flow-on benefits to other infectious diseases such as seasonal cold and influenza following non-pharmaceutical interventions.

Far-UVC represents a novel component for pandemic preparedness and response. As part of a comprehensive approach, far-UVC may come to complement other public health measures in prevention, mitigation, and response efforts to disease outbreaks.

## Methods

In this cost benefit study, we focussed on three specific environments: restaurants, offices, and waiting rooms in hospitals/doctors’ offices. Our selection is based on the undesirability or impracticality of mask usage in restaurants and offices, and the higher likelihood of contagiousness and more vulnerable individuals present in medical waiting rooms. Also considered was the frequency a standard person would come into contact with the spaces, thus we avoided custodial settings, for instance, which also had very high transmission risk [37, 38]. We excluded schools, despite their documented high incidence during initial epidemic phases, as we thought it was unlikely schools would be the first places to trial a new technology because of the increased vulnerability of children and inability to provide consent [39, 40]. These settings are in line with recommendations by the Centres for Disease Control and Prevention for selection of locations for upper-room ultraviolet germicidal irradiation, and priority spaces identified in the literature, which are extrapolated to use-cases for far-UVC [41].

To model the impact of introducing far-UVC lamps, we defined specific parameters for each environment, as described in *Appendix A*. These parameters include room size, number of occupants, number of infected occupants, duration of stay, and activity level [3]. The parameters were based on a combination of published literature (such as time use surveys) [42]. The number of infected people was varied to reflect different levels of population incidence based on Robert Koch Institute’s COVID and influenza reports. Additionally, we assumed a general ventilation rate of five air changes per hour (ACH) in every room, representing a blend of mechanically ventilated and naturally ventilated rooms [43]. This assumption is limited, however, by the few existing studies about the extent of ventilation in existing buildings.

The chosen settings try to depict a wide variety of the most common restaurants, offices and waiting rooms. Regarding restaurants we modeled a regular midsize restaurant (100 m²), an Irish pub (120 m²), a big fast food chain restaurant (180 m²) as well as a queue, where people would wait to get a takeaway lunch. For the waiting rooms, we selected general practitioner and clinical waiting rooms as well as an emergency department waiting room. The office scenarios represent a small (20 m², 3 desks), midsize (50 m², 8 desks) and large open floor office (100 m², 20 desks). All our chosen examples are based on reference places, personal measurements, or expert knowledge. The number of UV lamps was adjusted relative to the room volume to give recommended coverage as per the published literature. The lamp details used for modelling correspond to currently available 110 mW Krypton Chloride Excimer lamps.

Using the CERN Airborne Model for Indoor Risk Assessment (CAiMIRA21), we evaluate the expected changes in infection risk by the introduction of far-UVC lamps while ensuring they remain within the allowed exposure limit of 8 hours. CAiMIRA is a model of airborne pathogen transmission in indoor settings to quantify the risk of long-range airborne spread of SARS-CoV-2 virus, enabling the assessment of protective measures in the workplace. Since its inception it has been adapted to now power the World Health Organisation’s Indoor Airborne Risk Assessment (ARIA) tool [44].

We defined the prevalence of infectious individuals in indoor settings based on community incidence rates of respiratory diseases from the Robert Koch Institute. We considered three scenarios: a normal winter with mainly influenza and common colds lasting 22 weeks, a COVID-19-like pandemic with a wave lasting 4 weeks, and a more severe pandemic with a wave duration of 8 weeks, based on epidemiological studies of respiratory disease incidence spikes.

The efficacy of the lamps, characterized by the inactivation constant I], is translated into equivalent air changes per hour (eACH) based on empirical data presented by Eadie et al.[10]. For scenarios including (natural) ventilation and far-UVC lamps, the air-changes per hour are summed. Using CAiMIRA, we determine the difference in infections with and without far-UVC lamps, computing the number of avoided infections in each setting per scenario of infectious individuals in space. Our analysis further estimated the monetary benefits of avoided infections, including fewer hospitalizations, reduced sick days, and lower mortality.

These benefits are categorized into health care cost savings, economic productivity gains and monetized health benefits (intangible benefits to quality-of-life years QALY). The health care cost savings were calculated by multiplying the hospitalization rate by the average cost of a hospital stay. The economic productivity gains were calculated by multiplying the loss of productivity per sick day by the average number of sick days in each scenario. The monetized health benefits are calculated by multiplying the average QALY loss per infection by the QALY value given by The Swiss National COVID-19 Science Task Force [45]. Individual contributions of each as they relate to compounded calculations (e.g. total benefits) can be found in the supplementary materials.

Health and economic costs and benefits are informed by the literature and expert opinion. Using this comprehensive approach, we assess the potential health and economic impact of introducing far-UVC lamps in various indoor settings.

The expected costs for far-UVC lamps vary, depending on brand and availability. To model the estimated costs in our analysis we assume installation costs of CHF 1,350 per lamp and a power input of 11 W at an average life expectancy of 15,000 h or 10 years if used infrequently. For a lamp that is used for 8 h a day, 5 days a week, 22 weeks a year (for example, in an office) this would translate into annual costs of approximately CHF 170. It must be noted that the relatively high acquisition costs were divided over the lifetime of far-UVC lamps. QALY costs incurred from potential harmful effects as outlined in *Safety* have not been included here as there is limited published literature.

We also look at the cost for HEPA-filtration devices with typical costs of $540 per device, 60 W power input and a life expectancy of 3 years (Phillips 4000i air purifier is used as the example). Using a filter for 8 hours a day, 5 days a week, 22 weeks a year, this would translate into annual costs of roughly CHF 200. Again, the acquisition costs must be divided over the lifetime of HEPA-filtration devices. In the following years, prices of these devices will decrease, as only the filters require replacement, which cost approximately USD 90 (again, based on the cost of a Phillips replacement filter).

In summary, the cost of far-UVC lamps and HEPA-filters are similar, only varying slightly in our selected settings (±20%). With an increase in market size the price of far-UVC lamps is expected to go down [personal communication]. Technological improvements may further decrease the price for far-UVC lamps, which would make them more affordable in the long run due to lower energy consumption compared to air filters.

## Results

Results are presented by setting and outcome, and by some composite measures such as the benefit-cost ratio and the estimated benefits per capita.

### Restaurants

During a normal winter, the effect of far-UVC installation in restaurants positively affects economic productivity the most, with the benefits stemming from avoiding mild infections and reducing sick days. As expected, the cost benefit ratio increases dramatically with the severity of a pandemic.

During a pandemic, the greatest benefits are attributable to avoiding QALY-losses. In a severe pandemic wave compared to a COVID-19-like pandemic wave, a tenfold increase in financial savings are seen for the health care system and the national economy too.

We studied other types of hospitality settings including a pub and a large fast-food restaurant with both a queue for take-away customers and a seating area for guests eating in. The benefits division is very similar to the medium-sized restaurant.

It is postulated that restaurants achieve elevated benefits due to frequent interactions among diverse individuals over extended periods, thereby enhancing pathogen transmission in these environments.

**Table 1:** Benefits in CHF for the healthcare system in Switzerland, the national economy through avoided sick days and monetised health benefits from avoided infections by having far-UVC lamps installed in one medium-sized restaurant given three pandemic scenarios of varying severity.

| Benefits | Health Care Cost Savings | Economic Productivity Gains | Monetized Health Benefits |
| --- | --- | --- | --- |
| normal winter | 6'000 | 210'000 | 130'000 |
| COVID-19-like pandemic wave | 29'000 | 35'000 | 440'000 |
| severe pandemic wave | 330'000 | 310'000 | 23'000'000 |

**Table 2:** Benefits in EUR for the healthcare system in Germany, the national economy through avoided sick days and monetised health benefits from avoided infections by having far-UVC lamps installed in one medium-sized restaurant given three pandemic scenarios of varying severity.

| Benefits (restaurant) | Healthcare cost savings | Economic productivity gains | Monetized health benefits |
| --- | --- | --- | --- |
| normal winter | 2,000 | 70,000 | 60,000 |
| COVID-19-like pandemic wave | 10,000 | 12,000 | 200,000 |
| severe pandemic wave | 45,000 | 106,000 | 10,582,000 |

**Table 3:** Total benefits in CHF from avoided infections by having far-UVC lamps installed in one of the indicated types of restaurant given three pandemic scenarios of varying severity, in Switzerland.

| Sum of all benefits | Normal winter | COVID-19-like pandemic wave | Severe pandemic wave |
| --- | --- | --- | --- |
| Pub | 600'000 | 1'400'000 | 46'500'000 |
| Large fast food restaurant | 950'000 | 1'600'000 | 67'000'000 |
| Queue fast food restaurant | 52'000 | 87'000 | 3'700'000 |
| Medium-sized restaurant | 340'000 | 500'000 | 24'000'000 |

**Table 4:**
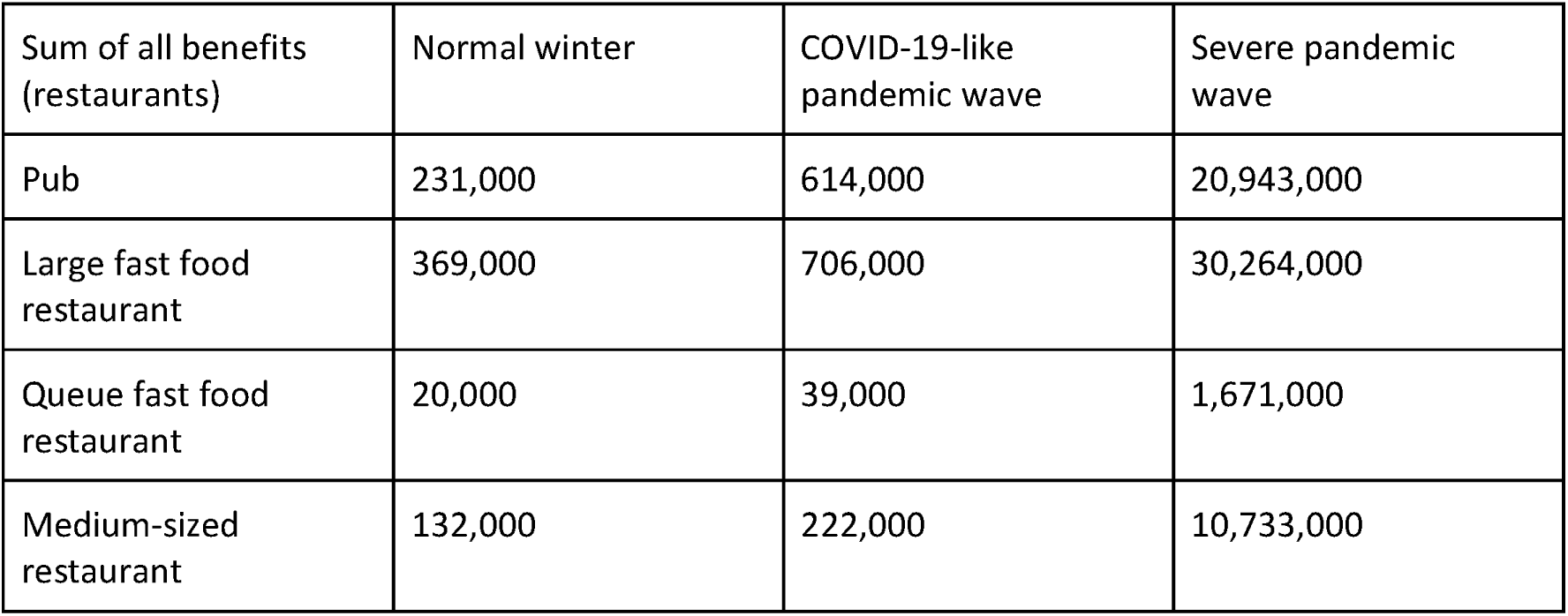
Total benefits in EUR from avoided infections by having far-UVC lamps installed in one of the indicated types of restaurant given three pandemic scenarios of varying severity, in Germany.

| Sum of all benefits (restaurants) | Normal winter | COVID-19-like pandemic wave | Severe pandemic wave |
| --- | --- | --- | --- |
| Pub | 231,000 | 614,000 | 20,943,000 |
| Large fast food restaurant | 369,000 | 706,000 | 30,264,000 |
| Queue fast food restaurant | 20,000 | 39,000 | 1,671,000 |
| Medium-sized restaurant | 132,000 | 222,000 | 10,733,000 |

### Waiting rooms

In total we considered four different waiting rooms. All scenarios were in a similar range for the number of avoided infections. Therefore, we decided to report the average over all scenarios. Like a moderately sized restaurant, national economic benefits matter most during a normal winter in a waiting room, whereas the share of monetized health benefits hugely increases with the severity of a pandemic. While during a normal winter most benefits stem from reducing sick days, during a pandemic much more benefits stem from the health benefits (added QALYs) (Figure 1). The health care cost savings and the productivity gains also increase eightfold in a severe pandemic compared to a COVID-19-like pandemic wave.

Although people visiting a doctor’s office or an emergency room are often suffering from acute illnesses, we refrained from adjusting the community incidence rate for a waiting room, since not every waiting room justifies a higher incidence rate, e.g. in a neurology ward. Consequently, this calculation likely underestimates the benefits of far-UVC lights in medical waiting rooms. Even though the waiting room scenario has a lower monetary benefit than the restaurant example, it should be considered for far-UVC lamp installation due to the vulnerable groups involved and the necessity of maintaining a functioning healthcare system during a pandemic.

**Table 5:** Benefits in CHF for the healthcare system, the national economy through avoided sick days and monetised health benefits from avoided infections by having far-UVC lamps installed in an average medical waiting room given three pandemic scenarios of varying severity, in Switzerland.

| **Average**<br>**Waiting Room** | **Health Care Cost**<br>**Savings** | **Economic**<br>**Productivity Gains** | **Monetized Health**<br>**Benefits** |
| --- | --- | --- | --- |
| normal winter | 290 | 10'000 | 6'500 |
| COVID-19-like<br>pandemic wave | 1'700 | 2'100 | 26'000 |
| severe pandemic wave | 17'000 | 16'000 | 1'200'000 |

**Table 6:** Benefits in EUR for the healthcare system, the national economy through avoided sick days and monetised health benefits from avoided infections by having far-UVC lamps installed in an average medical waiting room given three pandemic scenarios of varying severity, in Germany.

| Benefits (waiting room) | Healthcare cost savings | Economic productivity gains | Monetized health benefits |
| --- | --- | --- | --- |
| Normal winter | 100 | 3,500 | 3,000 |
| COVID-19-like pandemic wave | 600 | 700 | 12,000 |
| Severe pandemic wave | 2,300 | 5,500 | 553,600 |

**Fig.1.**
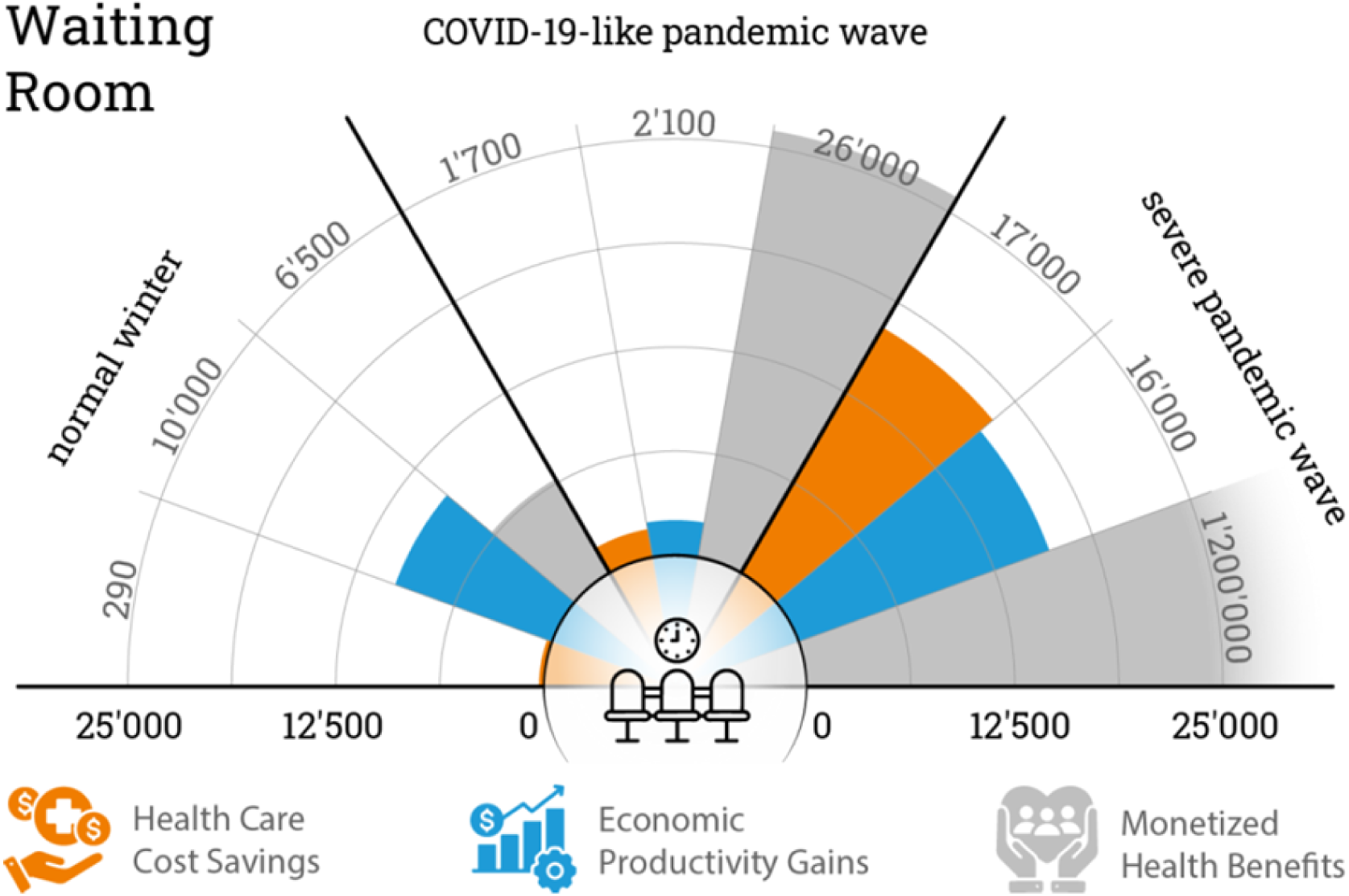
Overview of benefits gained through the implementation of far-UVC lamps in the indicated pandemic scenarios in an average medical waiting room in Switzerland. Health care cost savings are shown in orange, economic productivity gains in blue and monetized health benefits in grey

### Offices

Just like the restaurant and waiting room settings, productivity sees the greatest share of benefits during a normal winter in office spaces, whereas the share of monetized health benefits hugely increases with the severity of a pandemic. During a normal winter, most benefits stem from reducing sick days, while during a pandemic, considerably more benefits stem from monetized health benefits (Figure 2).

**Fig. 2.**
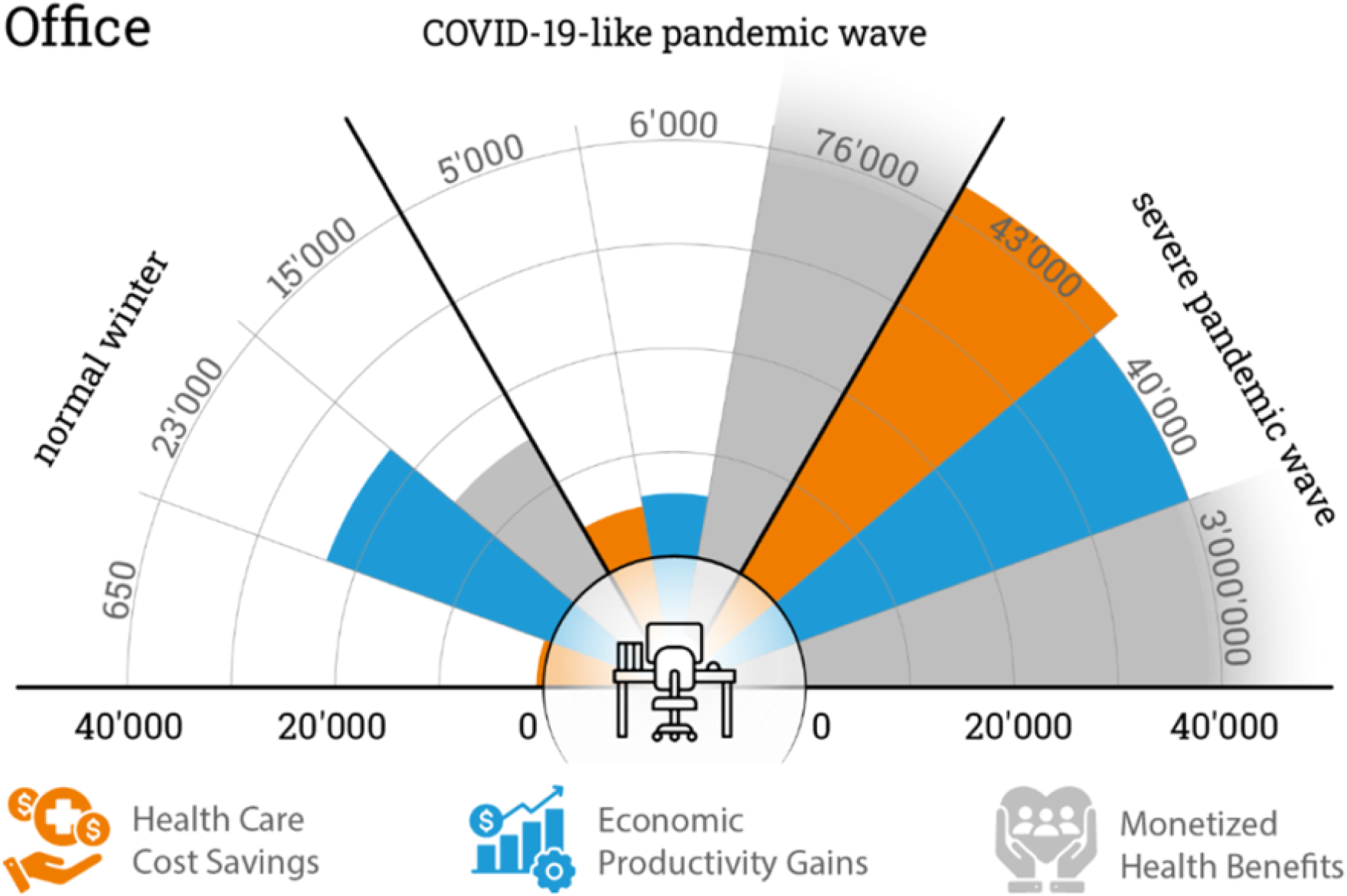
Overview of benefits gained through the implementation of far-UVC lamps in the indicated pandemic scenarios in a large office (100m²). Health care cost savings are shown in orange, economic productivity gains in blue and monetized health benefits in grey

In comparison to restaurants and waiting rooms, the importance of far-UVC lamps in offices increases with the number of people in the office, such that the benefits are relatively small in a three-person office, while the large office scenario shows substantial health and monetary benefits of installing far-UVC lamps (see Table 9).

**Table 7:**
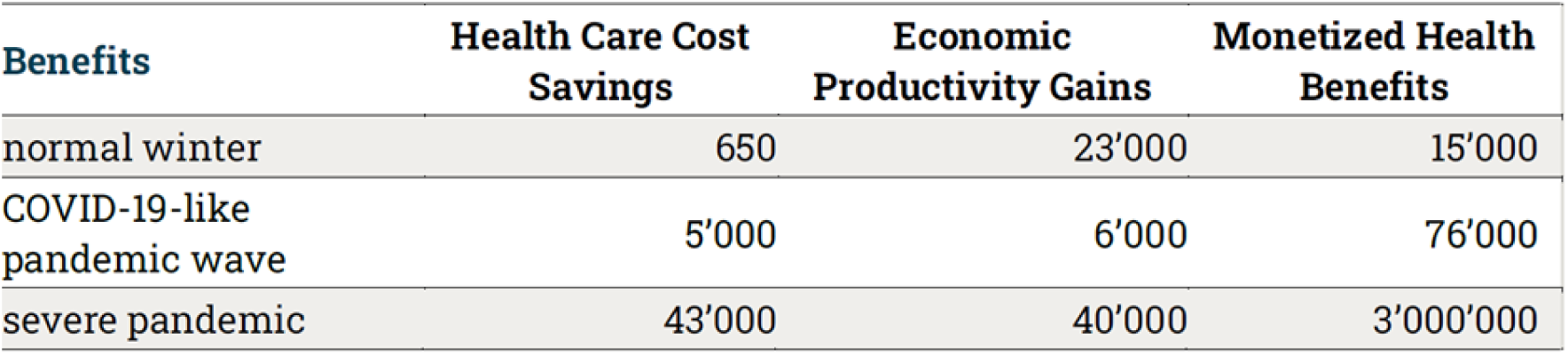
Benefits in CHF for the healthcare system, the national economy through avoided sick days and monetized health benefits from avoided infections by having far-UVC lamps installed in a large office (approx. 100m2, 20 desks) given three pandemic scenarios of varying severity.

| Benefits | Health Care Cost Savings | Economic Productivity Gains | Monetized Health Benefits |
| --- | --- | --- | --- |
| normal winter | 650 | 23'000 | 15'000 |
| COVID-19-like pandemic wave | 5'000 | 6'000 | 76'000 |
| severe pandemic | 43'000 | 40'000 | 3'000'000 |

**Table 8:** Benefits in EUR for the healthcare system, the national economy through avoided sick days and monetized health benefits from avoided infections by having far-UVC lamps installed in a large office (approx. 100m2, 20 desks) given three pandemic scenarios of varying severity, in Germany.

| Benefits (office) | Healthcare cost savings | Economic productivity gains | Monetized health benefits |
| --- | --- | --- | --- |
| Normal winter | 220 | 8,000 | 6,800 |
| COVID-19-like pandemic wave | 1,800 | 2,100 | 34,800 |
| Severe pandemic wave | 5,900 | 14,000 | 1,390,000 |

**Table 9:** Total benefits in CHF from avoided infections by having far-UVC lamps installed in the indicated type of office given three pandemic scenarios of varying severity, in Switzerland.

| Sum of all benefits in CHF | Normal winter | COVID-19-like pandemic wave | Severe pandemic wave |
| --- | --- | --- | --- |
| small office | 800 | 2'200 | 69'000 |
| medium office | 9'100 | 20'000 | 870'000 |
| large office | 39'000 | 87'000 | 3'100'000 |

### Cost benefit ratios

The benefit-to-cost ratio for far-UVC lamp installation is favorable across all settings, as indicated in Table 10 (Germany) and Table 11 (Switzerland). When accounting solely for the health care cost savings and productivity gains, excluding monetized health benefits, the cost-benefit ratio remains favorable across all scenarios. This indicates that far-UVC lamps are a cost-saving technology for the Swiss and German societies as studied herein. These findings may be extrapolated for other Western societies with similar lifestyles both in an average winter and a more severe pandemic scenario.

**Table 10:** Benefits to costs ratio for installing far-UVC lamps in the indicated settings in Germany. Benefits are summed over all three types and divided by the annual cost of the installed lamps. Darker shades of blue indicate higher benefits per EUR spent.

|  |  |  |  |
| --- | --- | --- | --- |
| Benefit/cost ratio | Restaurant | Waiting room | Office |
| Normal winter | 226 | 57 | 7 |
| COVID-19-like pandemic wave | 449 | 118 | 195 |
| Severe pandemic wave | 18,946 | 4,863 | 659 |

**Table 11:** Benefits to costs ratio for installing far-UVC lamps in the indicated settings in Switzerland. Benefits are summed over all three types and divided by the annual cost of the installed lamps. Darker shades of blue indicate higher benefits per CHF spent.

| **Benefits / cost ratio** | **restaurant** | **waiting room** | **office** |
| --- | --- | --- | --- |
| **normal winter** | 290 | 100 | 30 |
| **COVID-19-like pandemic wave** | 430 | 170 | 65 |
| **severe pandemic wave** | 20'500 | 6'700 | 2'300 |

In Switzerland, cost-benefit ratios ranged from one franc to: 30-290 CHF during a normal winter; 65- 430 CHF during a COVID-like pandemic; and 2,300-20,500 CHF during a severe pandemic (ranges correspond to different settings). In Germany, cost benefit ratios ranged from 1 euro to: 7-226 EUR during a normal winter; 118-449 EUR during a COVID-like pandemic; and 659-18,946 EUR during a severe pandemic. The large difference in cost benefit ratio between the countries may be attributed to not considering purchasing power parity in the calculations.

Another measure of the cost-effectiveness of health-related interventions is the incremental cost-effectiveness ratio (ICER). In this calculation, the status quo (ventilation at 5 ACH, no far-UVC lighting) is compared to the same room with far-UVC lighting installed. This comprised the incurred costs of implementing far-UVC lamps in the respective setting and the number of Quality-Adjusted Life Years (QALYs) saved through these far-UVC lamps. As described in *Table 12*, the incremental cost of saving one QALY is lowest in a restaurant during a severe pandemic, at 10 CHF per QALY. It is highest in an office/waiting room during a normal winter, at 1600 CHF per QALY. While Switzerland does not have an official willingness to pay threshold, this cost-effectiveness return is well within the USA and UK’s ICER thresholds, and the Swiss case law ruling of 100,000 CHF per QALY used in previous cost-effectiveness studies [46, 47]. Note that the ICER as presented here only considers the life years saved as benefits from the intervention, other benefits like avoided costs for the healthcare system or avoided sick days have not been included in the ICER.

**Table 12:**
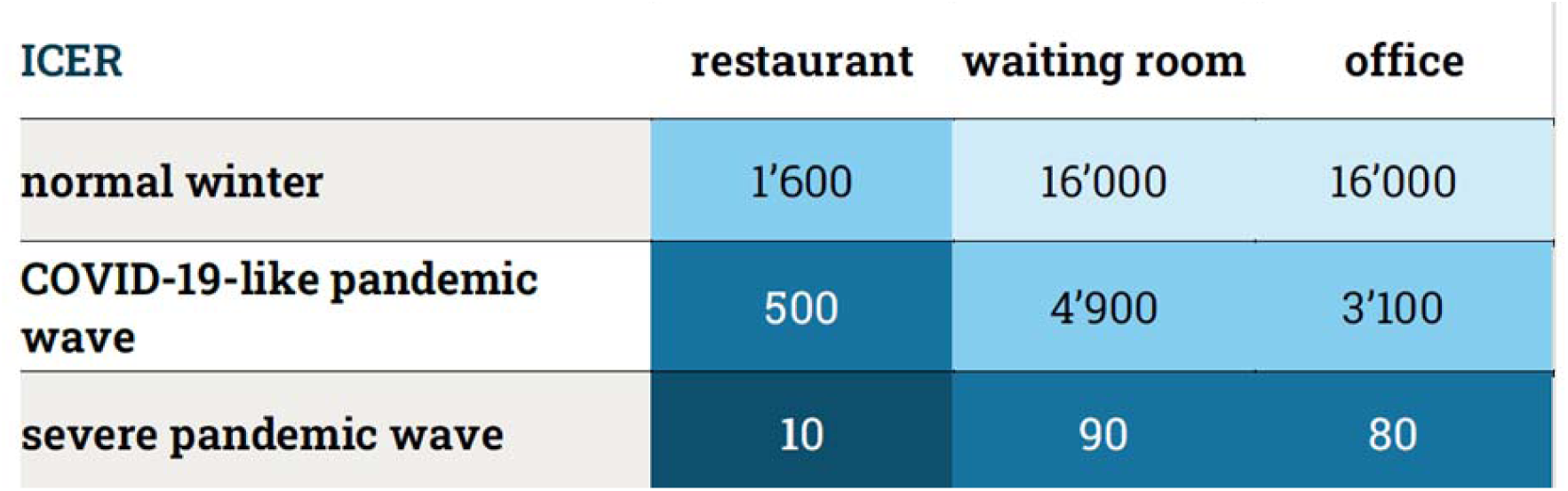
Incremental cost-effectiveness ratio (ICER) in CHF per QALY saved for installing far-UVC lamps in the indicated settings in Switzerland. Darker shades of blue indicate lower costs necessary to save a QALY.

In Germany and Switzerland, the greatest benefit is demonstrated in restaurants during a severe pandemic wave. As expected, the benefit:cost ratio decreases along with the severity of the pandemic. Nonetheless, even the smallest cost:benefit ratio, an office during a normal winter season, has a benefit that is estimated to be seven times the cost of the far-UVC lights.

If restaurants are analysed as the different types in our different scenarios, the greatest benefit accrues to fast food restaurants, followed by pubs/nightclubs. Our modelling demonstrated that benefit:cost ratio improves with the size of the office, with the smallest office included in Germany having a benefit of two times the cost of the lights, while the largest has an eleven times benefit. There was no discernible pattern between the different waiting rooms included.

### Per capita benefits

In addition, we calculated societal gains, i.e. total benefits minus costs, and avoided sick days per person annually if far-UVC was implemented in all restaurants, medical waiting rooms and offices in Switzerland. This was performed based on statistics about the number of each within the country and the existing estimations for the number of people visiting each during opening hours. Details can be found within the supplementary materials.

In 2022, in Switzerland, the average number of workdays missed by full-time employees amounted to 9.3 days [48]. Among these absences, approximately 69% were attributable to illness or accidents [48]. Therefore, a reduction of 2.4 days would signify a substantial decrease of about 37% in health-related absences or 26% in total work absences.

The gains per capita correspond to country-wide gains of CHF 12 billion in a normal winter, CHF 18 billion in a severe pandemic and CHF 860 billion in a severe pandemic. These comparatively high numbers, especially in the severe pandemic scenario need to be assessed with caution as we do not account for infections occurring in settings other than restaurants, offices and waiting rooms. Especially in scenarios with very high incidences among the population this assumption becomes increasingly distorting and will lead to an overestimation of our results. Nonetheless, the societal gains in a severe pandemic will be driven by very high numbers of avoided deaths and thus are plausibly very high compared to, e.g., the Swiss Gross Domestic Product.

Our findings highlight the importance and cost-effectiveness of improved indoor air quality during seasonal cold and flu waves, and how these increase during pandemics – particularly relevant given the threat of future pandemics and need for preparedness.

**Table 13:** Gains per Capita (= total benefits – costs) and annual avoided sick days per person due to introducing far-UVC in all restaurants, waiting rooms and offices in Switzerland.

| **Societal gains** | **gains (CHF) / capita** | **Annual avoided sick days / capita** |
| --- | --- | --- |
| **normal winter** | 1'400 | 2,4 |
| **COVID-19-like pandemic wave** | 2'100 | 0,4 |
| **severe pandemic** | 98'000 | 3,7 |

### Comparison to other indoor air quality measures

The intended usage of filters and lamps are similar, making costs and benefits readily comparable. We found very similar costs associated with both HEPA and far-UVC. Both rely on the assumption of a homogeneous mixture in the room. HEPA also filters out nanoparticles but does not reduce the concentration of ozone, CO2 and other toxic chemicals by themselves. For this, additional activated carbon filters or ventilation is necessary. From an end-user perspective, a large difference stems from much higher noise levels generated by HEPA filtration especially at high clean air delivery rates compared to the silent far-UVC light emission. Furthermore, HEPA filtration requires considerably more energy (60 W per filter vs 11 W per far-UVC device). Thus, far-UVC could contribute to a more sustainable building footprint.

In our calculations, we have assumed far-UVC’s efficacy based on a study by Eadie et al on *Staphylococcus Aureus* bacteria. Some studies suggest far-UVC lamps are 10-100 times more effective than this due to the varying effects of far-UVC light on different viruses and bacteria [18, 49]. It appears that especially for SARS-CoV-2 and influenza-like viruses far-UVC light is highly effective [18, 49]. Further research is required, but early literature has shown promising results. If the lamps are at least 10 times more effective than assumed in this study, they will consequently be much more cost efficient than HEPA filters and plausibly reach levels of virus inactivation that are unobtainable through HEPA filter placements.

## Discussion

A cost-benefit analysis of far-UVC light in suppressing infectious diseases is multifaceted and complex. Assumptions were used as a necessity. This discussion attempts to underline sources of uncertainty within the study.

The study hinges on plausible assumptions about far-UVC lamps’ effectiveness in varied environments (see *Methods*) with current infection rates and lamp efficacy estimates grounded in recent research, which may need to be updated as research evolves.

The modelling of avoided infections depends on the dynamics of specific pathogens and their transmission pathways. Changes in disease transmission patterns, caused by new variants or changes in human behaviour, could influence the study results [13]. The study is based on data available at the time of the analysis. Changes in technology, cost structures, or disease patterns can affect the relevance of the results over time. For instance,

⍰ The air changes per hour are based on current EU guidelines and may not reflect the reality of ventilation in different settings, particularly older buildings.
⍰ During a normal winter we assume use only during the six coolest months of the year, as these tend to have the highest rates of respiratory illness.
⍰ COVID-like pandemic assumes a 4-week peak based on the Omicron peak in Switzerland. This assumption partly explains why benefits are significantly lower than other scenarios.

Benefits were split into savings to the health care system, productivity gains and monetised health benefits (i.e. QALY gains) - we chose to count these separately so that decision makers can extract which is most relevant to them. For instance, business owners considering implementing UVC in offices might be most interested in productivity gains.

A limitation of this study is the lack of accounting for potential disease transmission in locations beyond the spaces equipped with far-UVC sources, as well as the exclusion of any mitigated transmission risk from individuals visiting far-UVC-equipped areas. The interplay of both effects is hard to assess and has thus been excluded here. Note that this is likely an underestimation of the total benefits in low incidence scenarios, as we did not account for the multiplier effect of avoided infections.

The potential health impacts of far-UVC on humans and animals are the subject of ongoing research. The ecological costs, including the electricity used and the disposal of far-UVC lamps, could also be considered, but were excluded from this study. Costs of potential harmful effects as outlined in *Safety* have not been included.

Far-UVC is part of an array of approaches enhancing indoor air quality. A holistic approach including ventilation and filtration is likely to be more effective than the singular use of far-UVC, as is demonstrated in the additive benefits of ventilation, HEPA filtration and far-UVC lights in our scenarios.

The effectiveness of far-UVC lamps is dependent on their correct installation and maintenance, though this is much less prone to human error compared to masks or manual disinfection. Errors in implementation could reduce the expected benefits.

Public perception and acceptance of far-UVC technologies will be an important factor for successful implementation. Public engagement may need to be factored into costs.

The effectiveness of far-UVC lamps varies depending on the type of premises (e.g., size, ventilation, foot traffic) in which they are installed. The most promising environments for the installation may be identified based on the results of this study.

The decision on where to install far-UVC lamps raises questions of equity and access, especially in relation to resource-poor contexts.

### Policy recommendations and next steps

Given the cost efficiency of far-UVC lamps, the following recommendations are made to support public health and pandemic preparedness. However, the implications of implementation will require ongoing evaluation.

#### Private Sector

1. Enhance Infection Protection through Monitoring and Air Quality Improvement

Regular air quality measurements and installation of systems to manage indoor air are recommended for the private sector. This helps businesses ensure safer environments and promote well-being of customers and employees. Far-UVC can contribute to the goal of decreasing disease transmission if implemented within the current European guidelines and in combination with ventilation. This is hereby demonstrated to be cost-effective, and to reduce sick days.

#### Public Institutions

2. Conduct real-world and feasibility studies of far-UVC

Real-world studies are necessary to assess the practical applicability of far-UVC light. Research could be commissioned by public health offices or national research programs.

3. Monitor indoor air quality and ensure transparency

Comprehensive data on indoor air quality is lacking in countries like Germany and Switzerland. Surveys and public reporting can identify problem areas and define targeted measures for improvement, such as ventilation.

4. Anchor the state’s position as a role model

By setting air quality standards for public buildings, the government promotes public health and encourages other sectors.

5. Create incentives to improve indoor air quality

Financial incentives can help overcome implementation barriers and play a significant role in encouraging implementation of public health measures. Far-UVC may also contribute to achieving certifications like Leadership in Energy and Environmental Design.

6. Equip critical infrastructure with far-UVC technology for pandemic response

Facilities critical during pandemics, such as hospitals and government buildings, should consider long-term far-UVC installations as part of their pandemic response plans.

## Conclusions

Far-UVC technology can offer a more reliable and less intrusive solution for mitigating indoor disease transmission across office, restaurant and waiting room settings. Integrating far-UVC into public health strategies can enhance community resilience against infectious diseases. Far-UVC can significantly reduce health care costs by reducing infections, avoiding hospitalizations and reducing the need for expensive treatments. It also provides economic benefits through productivity gains due to fewer sickness-related absences of employees. Initial research appears promising, and we encourage real-world trials of this technology to report findings.

## Supporting information

Setting assumptions used in the research

## Data Availability

All data produced in the present work are contained in the appendix through our "setting assumptions" table

## Acknowledgements

We thank the following reviewers for their time and expertise:

Dr. Nicolas Banholzer, Postdoctoral Research Fellow, Institute of Social and Preventive Medicine, University of Bern

Dr. Richard Bruns, Senior Scholar and Economist, Johns Hopkins Center for Health Security

Dr. Jasper Götting, Virologist, Convergent Research

Jakob Graabak, Foresight Director, International Center for Future Generations

Harry Koos, PhD student in Health Policy / Health Economics, Stanford School of Medicine

Dr. Michael Riediker, Director, Swiss Centre for Occupational and Environmental Health

Dr. Lisa Fischer, d-fine

Dr. Robert Görke, d-fine, Partner Pharma and Healthcare

Dr. Daniel Staudenmann, MD, Cantonal Hospital of Aarau

Prof. Dr. Ernest Weingartner, Lecturer in Sensor and Aerosol Technology, University of Applied Sciences and Arts Northwestern Switzerland

Richard Williamson, Program Director far-UVC, Blueprint Biosecurity

d-fine is a European consulting firm which, by means of scientifically minded employees, provides innovative and future-proof solutions through sustainable technological implementation. d-fine supported the project as part of a pro bono engagement. The current lack of cost benefit studies and the organisation’s interest in healthcare and technology prompted d-fine to support the project with their expertise.

www.d-fine.com

Pour Demain is an independent European think tank working towards a safe and positive future. As a nonprofit organisation Pour Demain is committed to epidemic and pandemic preparedness. Innovations such as far-UVC represent possibly highly effective interventions to reduce the impact of future pandemics, and could save many lives. Pour Demain is dedicated to evidence-based policies.

www.pourdemain.ngo

## Appendix A: Table of Assumptions

| Parameter | Value | Source |
| --- | --- | --- |
| *COVID-19-LIKE PANDEMIC WAVE* |  |  |
| Symptomatic COVID-19 case | 75% | [50, 51, 52] |
| Long COVID | 10% | [53] |
| Hospitalization rate overall | 10% | [54] |
| Mortality COVID-19 overall | 4% | [55] |
| Cost of hospitalization | 30'250 CHF | [56] |
| Cost of disability case for healthcare system | 8'500 CHF | [56] |
| Sick days per symptomatic COVID-19 infection | 5 | [56] |
| Sick days per COVID-19 hospitalization | 15 | [57, 58, 59] |
| Sick days per Long Covid case | 84 | [60] |
| QALYs lost per death due to COVID-19 | 11 | [61] |
| Losses of QALYs associated with symptomatic-COVID-19 case | 0,008 | [62] |
| Losses of QALYs associated with a long COVID infection | 0,034 | [62] |
| Losses of QALYs associated with a COVID-19 hospitalization | 0,020 | [62] |
| *NORMAL WINTER* |  |  |
| Hospitalization rate overall | 0,30% | Calculated from [63] |
| Mortality overall | 0,0060% | [64] |
| Cost of hospitalization | 10'292 CHF | [65] |
| Sick days per infection | 3 | [66] |
| Sick days per hospitalization | 17 | Assumption, similar to covid (20 sick days in total) |
| QALYs lost per death due to resp infection | 3 | Estimation based on [67] |
| Losses of QALYs associated with a respiratory infection | 0,0038 | [68] |
| Losses of QALYs associated with hospitalization | 0,0118 | [68] |
| *SEVERE PANDEMIC WAVE* |  |  |
| disability induced | 10% | Assumption to model a more severe pathogen than covid-19 |
| Hospitalization rate overall | 30% | Assumption to model a more severe pathogen than covid-19 |
| Mortality overall | 20% | Assumption to model a more severe pathogen than covid-19 |
| Cost of hospitalization | 30.250 CHF | As for covid-like pandemic |
| Cost of disability case for healthcare | 8.500 CHF | As for covid-like pandemic |
| Sick days per infection | 10 | Assumption |
| Sick days per hospitalization | 20 | Assumption |
| Sick days per case of disability | 100 | Assumption |
| QALYs lost per death | 20 | Assumption |
| Losses of QALYs associated with an infection | 0,01 | Assumption extrapolated from covid and influenza |
| Losses of QALYs associated with a disability induced from Infection | 0,05 | Assumption extrapolated from covid and influenza |
| Losses of QALYs associated with hospitalization | 0,04 | Assumption extrapolated from covid and influenza |
| *GENERAL ASSUMPTIONS* |  |  |
| Loss of productivity per sick day | 360 CHF | [56] |
| Economic equivalent of 1 QALY | 175'000 CHF | [56] |

## Contribution

Conceptualization, Data curation, Methodology, Project administration, Funding Acquisition, Writing – original draft

Investigation, Writing – original draft, review & editing

Data curation, Formal analysis, Investigation, Methodology, Validation, Writing – original draft, Writing – review & editing

Project administration, Writing – original draft

Data curation, Formal analysis, Writing review & editing

Writing – review & editing, Formal analysis

Formal analysis, Writing – review & editing

Data curation, Formal analysis

